# Trends in mortality of alcoholic liver disease among adults in the United States, 1999-2017

**DOI:** 10.1101/2020.06.17.20133827

**Authors:** Emily Ryu, Harry H. Xia, Grace L Guo, Lanjing Zhang

**Author notes:** Correspondence: Lanjing Zhang, MD, Department of Pathology, Princeton Medical Center, 1 Plainsboro Rd., Plainsboro, NJ 08563.

## Abstract

Some subtypes of alcoholic liver disease (ALD) recently had increasing prevalence or mortality. Prevalence of alcoholic fatty liver disease was increased. Mortality of alcoholic hepatitis and cirrhosis also had upward trends. However, trends in ALD- mortality and related factors are unclear. We therefore examined trends in age-standardized ALD-mortality among U.S. adults by factors using multivariable piecewise log-linear models. We collected mortality-data (age-standardized for the 2000 U.S. standard population) from the Centers for Disease Control and Prevention Wide-ranging Online Data for Epidemiologic Research database (CDC WONDER), using the Multiple Cause of Death Data to identify all ALD deaths in the United States for 1999-2017. We identified 296,194 deaths of ALD during 1999-2017. Trends in multivariable-adjusted, age-standardized mortality did not differ by sex, race, age or urbanization. The age-standardized mortality ratios of male/female, White/non-White and Metropolitan/Non-Metropolitan were 2.346, 1.657 and 0.851 in 2017, respectively. Strikingly, our multivariable model showed that subjects of 65+ years had the highest and the fastest growing mortality in the 3 age-groups. These findings highlight the continuation of health disparities in ALD, particularly in elderly subjects. Further works are warranted to validate and delineate the associated factors.

---

Likely due to increasing alcohol-use, some subtypes of alcoholic liver disease (ALD) recently had increasing prevalence or mortality. Prevalence of alcoholic fatty liver disease was increased.^1,2^ Mortality of alcoholic hepatitis and cirrhosis also had upward trends.^3,4^ However, trends in ALD-mortality and related factors are unclear. We therefore examined trends in age-standardized ALD-mortality among U.S. adults by factors using multivariable piecewise log-linear models.

## Methods

We collected mortality-data (age-standardized for the 2000 U.S. standard population) from the Centers for Disease Control and Prevention Wide-ranging Online Data for Epidemiologic Research database (CDC WONDER), using the Multiple Cause of Death Data to identify all ALD deaths in the United States for 1999-2017. Rates in WONDER were suppressed when the death count was <20. This study is exempt from the approval from an institutional review board (IRB) due to the use of de-identified publicly available data on the deceased U.S. residents.

Subjects were included if they were aged 25 years or older and had ALD as underlying death-cause on death certificates (International Classification of Diseases, code K70). Data were also extracted by the following variables: Sex (female/male), 20-year age group (25-44 years, 45-64 years, and 65+ years), race (White/non-White), urbanization (metro/non-metro, based on the 2006 NCHS Urban-Rural Scheme for Counties). An Institutional Review Board approval was not required because this was not human-subject research (all subjects deceased at the time of study) and we used de-identified, publicly available data.

Trends in mortality over time were explored using the National Cancer Institute’s joinpoint regression software (version 4.7.0.0) to determine possible presence of 1 trend change-point. We then used multivariable piecewise log-linear models to adjust covariables (Stata, version 15). Specifically, trends in age-standardized mortality were further adjusted for sex, race (White/non-White), 3-tier age groups and urbanization, except when the variable was used as the major factor. The years with death-count fewer than 20 were excluded due to unreliable data. Based on the multivariable piecewise linear model and adjusted to sex, race and urbanization, we predicted the age-standardized mortality among U.S. adults during 1999-2040 by age-group, using the age-standardized mortality ratios of male/female White/non-White and Metropolitan/Non-Metropolitan in 2017. Given a recently-reported change-point of trend in ALD mortality during 2007-2017,^4^ we analyzed the trend and its potential change-point using a similar multivariable piece-wise log-linear model. All P values were 2-sided, and only *P*<0.05 was considered statistically significant.

## Results

We identified 296,194 deaths of ALD during 1999-2017. Trends in multivariable-adjusted, age-standardized mortality did not differ by sex, race, age or urbanization (**Table**). The age-standardized mortality ratios of male/female, White/non-White and Metropolitan/Non-Metropolitan were 2.346, 1.657 and 0.851 in 2017, respectively. Based on these ratios and models, we predicted the sex-, race- and urbanization-adjusted age-standardized mortality of ALD during 1999-2040 (**Figure**). No change-points were identified during 2007-2017 (P=0.482). Strikingly, our multivariable model showed that subjects of 65+ years had the highest and the fastest growing mortality in the 3 age-groups.

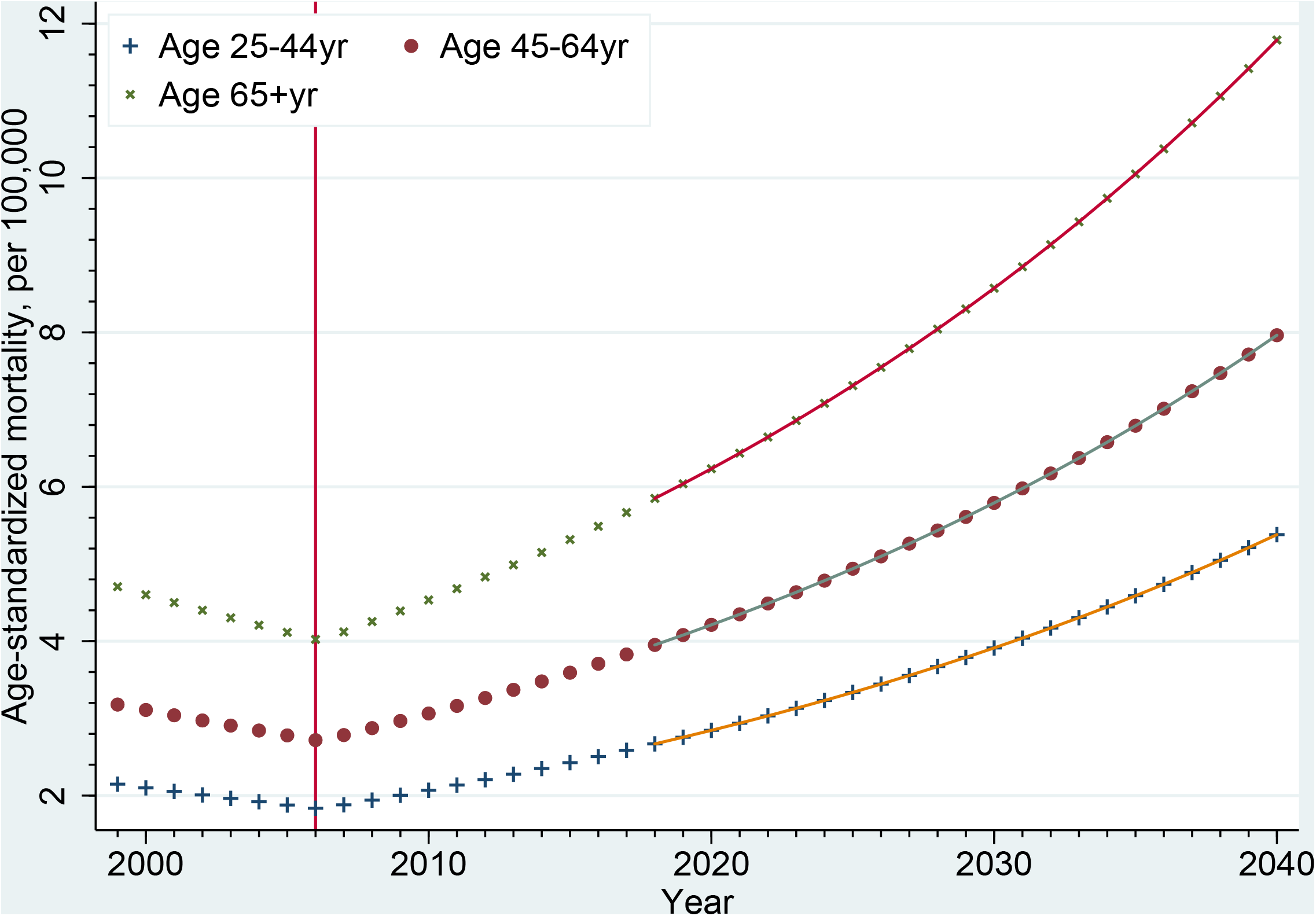
Trends in sex-, race- and urbanization-adjusted age-standardized mortality of alcoholic liver disease among U.S. adults by age group during 1999-2017 and beyond. There was a changing point of the trends in all age groups (vertical line, 2006) during 1999-2017. For data points in the years after 2017 (dash line), the ratios of male/female, white/non-white and metropolitan/non-metropolitan were assumed similar to those in 2017, and used for respective mortality prediction. Analysis limited to adults aged 25+ years.

**Table.**
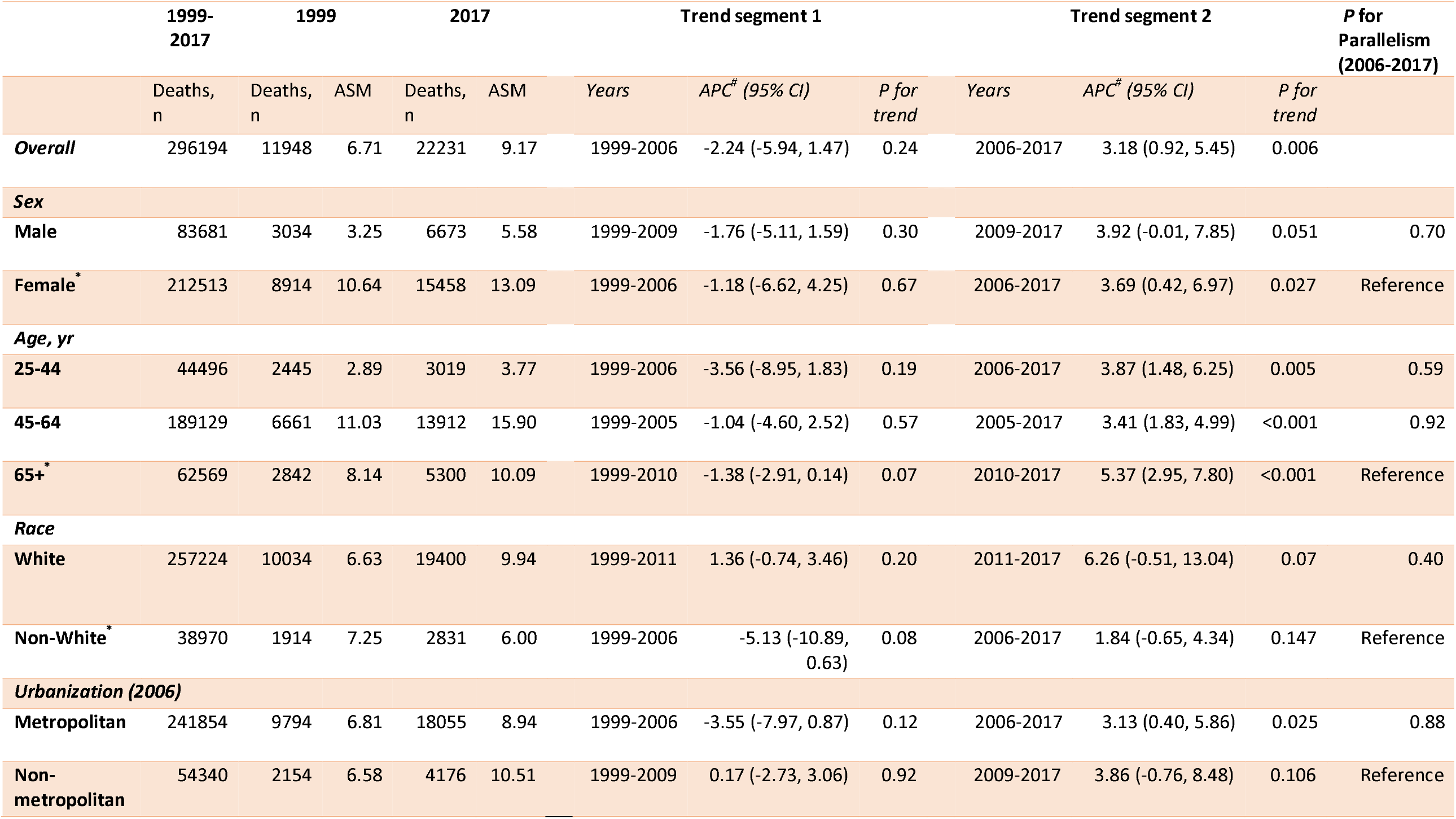
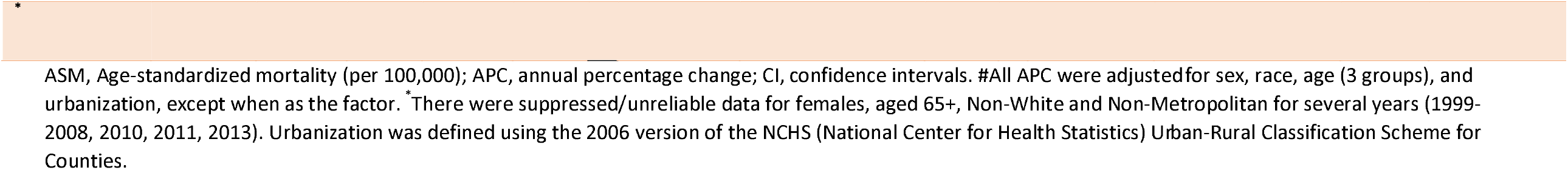
Multivariable-adjusted trends in age-standardized mortality of alcoholic liver disease among U.S. adults aged 25+ years, 1999-2017.

|  | 1999-2017 | 1999 |  | 2017 |  | Trend segment 1 |  |  | Trend segment 2 |  |  | P for Parallelism (2006-2017) |
| --- | --- | --- | --- | --- | --- | --- | --- | --- | --- | --- | --- | --- |
|  | Deaths, n | Deaths, n | ASM | Deaths, n | ASM | Years | APC<sup>#</sup> (95% CI) | P for trend | Years | APC<sup>#</sup> (95% CI) | P for trend |  |
| Overall | 296194 | 11948 | 6.71 | 22231 | 9.17 | 1999-2006 | -2.24 (-5.94, 1.47) | 0.24 | 2006-2017 | 3.18 (0.92, 5.45) | 0.006 |  |
| Sex |  |  |  |  |  |  |  |  |  |  |  |  |
| Male | 83681 | 3034 | 3.25 | 6673 | 5.58 | 1999-2009 | -1.76 (-5.11, 1.59) | 0.30 | 2009-2017 | 3.92 (-0.01, 7.85) | 0.051 | 0.70 |
| Female* | 212513 | 8914 | 10.64 | 15458 | 13.09 | 1999-2006 | -1.18 (-6.62, 4.25) | 0.67 | 2006-2017 | 3.69 (0.42, 6.97) | 0.027 | Reference |
| Age, yr |  |  |  |  |  |  |  |  |  |  |  |  |
| 25-44 | 44496 | 2445 | 2.89 | 3019 | 3.77 | 1999-2006 | -3.56 (-8.95, 1.83) | 0.19 | 2006-2017 | 3.87 (1.48, 6.25) | 0.005 | 0.59 |
| 45-64 | 189129 | 6661 | 11.03 | 13912 | 15.90 | 1999-2005 | -1.04 (-4.60, 2.52) | 0.57 | 2005-2017 | 3.41 (1.83, 4.99) | <0.001 | 0.92 |
| 65+* | 62569 | 2842 | 8.14 | 5300 | 10.09 | 1999-2010 | -1.38 (-2.91, 0.14) | 0.07 | 2010-2017 | 5.37 (2.95, 7.80) | <0.001 | Reference |
| Race |  |  |  |  |  |  |  |  |  |  |  |  |
| White | 257224 | 10034 | 6.63 | 19400 | 9.94 | 1999-2011 | 1.36 (-0.74, 3.46) | 0.20 | 2011-2017 | 6.26 (-0.51, 13.04) | 0.07 | 0.40 |
| Non-White* | 38970 | 1914 | 7.25 | 2831 | 6.00 | 1999-2006 | -5.13 (-10.89, 0.63) | 0.08 | 2006-2017 | 1.84 (-0.65, 4.34) | 0.147 | Reference |
| Urbanization (2006) |  |  |  |  |  |  |  |  |  |  |  |  |
| Metropolitan | 241854 | 9794 | 6.81 | 18055 | 8.94 | 1999-2006 | -3.55 (-7.97, 0.87) | 0.12 | 2006-2017 | 3.13 (0.40, 5.86) | 0.025 | 0.88 |
| Non-metropolitan | 54340 | 2154 | 6.58 | 4176 | 10.51 | 1999-2009 | 0.17 (-2.73, 3.06) | 0.92 | 2009-2017 | 3.86 (-0.76, 8.48) | 0.106 | Reference |

\*
ASM, Age-standardized mortality (per 100,000); APC, annual percentage change; CI, confidence intervals. #All APC were adjusted for sex, race, age (3 groups), and urbanization, except when as the factor. \* There were suppressed/unreliable data for females, aged 65+, Non-White and Non-Metropolitan for several years (1999-2008, 2010, 2011, 2013). Urbanization was defined using the 2006 version of the NCHS (National Center for Health Statistics) Urban-Rural Classification Scheme for Counties.

## Discussion

Age-standardized mortality due to ALD among U.S. adults decreased during 1999-2006, but continued increasing ever since. There were no significant differences in ALD-mortality trends by sex, race, age and urbanization. Therefore, the mortality disparities of ALD by these factors^3-5^ will continue increasing. These findings are thus alarming and highlight the need for awareness of ALD burdens and increasing health disparities.

Limitations include potential misclassification of causes on death certificates, which were used before^4,5^ and highly specific for underly causes of death other than pneumonia.^6^ Owing to long study-period, we might underestimate the number of trend change-point. To address this issue, we conducted a sensitivity analysis for 2007-2017, but did not identify any additional change-points, which were shown in a univariable model.^4^ The difference might be attributable to multivariable versus univariable model and aged 25+ versus 20+ years in this and the prior studies, respectively.

## Data Availability

All available at the CDC WONDER website.

## Author Contributions

Dr Zhang had full access to all of the data in the study and takes responsibility for the integrity of the data and the accuracy of the data analysis.

Concept and design: Ryu, Guo, Zhang.

Acquisition, analysis, or interpretation of data: Ryu, Xia, Zhang. Drafting of the manuscript: Ryu.

Critical revision of the manuscript for important intellectual content: All authors. Statistical analysis: Ryu, Zhang.

Supervision: Zhang.

